# Reliability of self-sampling for accurate assessment of respiratory virus viral and immunologic kinetics

**DOI:** 10.1101/2020.04.03.20051706

**Authors:** Alpana Waghmare, Elizabeth M. Krantz, Subhasish Baral, Emma Vasquez, Tillie Loeffelholz, E. Lisa Chung, Urvashi Pandey, Jane Kuypers, Elizabeth R Duke, Keith R. Jerome, Alexander L. Greninger, Daniel B. Reeves, Florian Hladik, E. Fabian Cardozo-Ojeda, Michael Boeckh, Joshua T. Schiffer

**Affiliations:** Vaccine and Infectious Diseases Division, Fred Hutchinson Cancer Research Center; Department of Pediatrics, University of Washington; Center for Clinical and Translational Research, Seattle Children’s Research Institute; Department of Obstetrics and Gynecology, University of Washington; Department of Laboratory Medicine, University of Washington; Department of Medicine, University of Washington; Clinical Research Division, Fred Hutchinson Cancer Research Center

## Abstract

The SARS-CoV-2 pandemic demonstrates the need for accurate and convenient approaches to diagnose and therapeutically monitor respiratory viral infections. We demonstrated that self-sampling with foam swabs is well-tolerated and provides quantitative viral output concordant with flocked swabs. Using longitudinal home-based self-sampling, we demonstrate nasal cytokine levels correlate and cluster according to immune cell of origin. Periods of stable viral loads are followed by rapid elimination, which could be coupled with cytokine expansion and contraction using mathematical models. Nasal foam swab self-sampling at home provides a precise, mechanistic readout of respiratory virus shedding and local immune responses.

---

The COVID-19 pandemic is an unprecedented event in modern history. As of March 30, there are 784,000 documented COVID-19 cases, which is surely an underestimation, and 37,638 deaths worldwide with rapidly expanding outbreaks ongoing in dozens of countries^1^. Morbidity and mortality rates are dangerously high in the elderly and those with medical comorbidities^2,3^. Current informal estimates suggest that 20-70% of humans may become infected without global deployment of a vaccine, which is unlikely to occur in the next year. While social distancing has proven effective in several countries in Asia, these measures might not be sustainable without crippling the global economy and may not be as successfully implemented elsewhere. Under optimistic projections, social distancing may push COVID-19 to a fluctuating pattern during which periodic outbreaks necessitate repeated implementation of social distancing^4^. In all likelihood, this highly contagious and lethal respiratory virus will likely circulate widely for years to come^5^.

A critical research priority is to develop rapid molecular tests that provide accurate diagnosis, determine infectiousness and transmissibility, and allow for monitoring of viral load during therapy^6^. For numerous viral infections, including influenza, viral load correlates with disease severity and secondary household attack rate^7-9^. Early studies suggest that peak viral load differentiates mild from severe COVID-19^10^.Furthermore, viral load monitoring during antiviral therapy is a mainstay for various human infections including HIV, hepatitis B, cytomegalovirus and hepatitis C infections^11-17^. Particularly for viruses such as SARS-CoV-2 for which severe clinical outcomes occur in a minority of patients, viral load may serve as a useful surrogate marker to design smaller, but still sufficiently powered treatment studies^4,10^.

Another major unmet medical need is the ability to frequently measure the local mucosal immune response during the course of infection. It is increasingly recognized that tissue resident T-cells and antigen presenting cells are phenotypically and functionally distinct from circulating immune cells, especially in the setting of respiratory viral infections^18-20^. Therefore, measuring immune cells in blood can fundamentally misclassify the agents responsible for viral elimination at the local level. To assess tissue resident immune cells requires biopsies which are difficult to obtain during active infection. Yet, important shifts in the immune response against respiratory viruses likely occur rapidly and in stages during the early and late phases of viral shedding^21^. Serial measurement of local cytokines may provide a window into the local cellular response^22^, but has yet to be validated from longitudinal clinical samples.

Self-testing for respiratory viruses has been promoted for more than a decade and successfully performed both in research and primary care settings, but regulatory agencies have been slow to accept patient collected samples as valid, especially in the home setting. Recently issued initial guidelines from the United States Food and Drug Administration (FDA) required nasopharyngeal (NP) sampling using flocked swabs for diagnosis of COVID-19 by clinical laboratories^23^. However, as the demand for testing exponentially increases, NP swab availability significantly hampers effective and efficient testing and identification of COVID-19-infected individuals. Currently licensed flocked swabs may not be optimal for patients with vulnerable mucosal membranes and low platelet counts (e.g. following cytotoxic chemotherapy) because they are associated with some discomfort and possible bleeding. Moreover, their general level of discomfort may deter participants from collecting longitudinal samples. This may limit widespread use for self-testing, especially as surveillance testing or for use in vulnerable patients or children. Importantly, a reliable and comfortable home-based self-testing methodology is needed to prevent potentially infected individuals from entering healthcare facilities to be tested and transmitting virus to healthcare workers and other patients. Initial data on foam swabs are promising, suggesting a broader role for home-based self-swabbing for respiratory viral pandemics^24,25^.

Here we report data on a novel respiratory virus detection method using self-collected nasal foam swabs. This methodology expands our testing armamentarium with easily collected and comfortable swabs that can be applied to viral load and cytokine kinetic studies. Most importantly, they can be easily scaled and used at home in this time of severe testing shortages and dangerous transmission risk.

## Results

### Concordance between foam and flocked nasal swabs for viral detection

Fifteen participants were enrolled within 3 days of respiratory symptom onset **(Supp Table 1)**. Four participants were negative for any respiratory virus from all swabs on our multiplex PCR panel **(Table 1)**. Participants swabbed each nostril with a foam swab and a flocked swab, randomized by order of swab type. Combining results from both nostrils, foam and flocked swabs were concordant for viral detection in 22/30 samples (73.3%). Among the 12 samples positive by flocked swab, 3 were negative by foam swab. Among 14 samples positive by foam swab, 5 were negative by flocked swab **(Supp Table 2)**. Discrepant results occurred exclusively in samples with low viral load (<4 log_10_ viral copies/mL) **(Table 1)**.

**Table 1:** Viral loads in matched foam versus flocked swabs in participants with new onset respiratory symptoms.

| **Participant number** | **Virus** | **Foam right Log10 copies/ml** | **Foam left Log10 copies/ml** | **Flocked right Log10 copies/ml** | **Flocked left Log10 copies/ml** |
| --- | --- | --- | --- | --- | --- |
| p1 | PIV3 | 6.12 | 3.79 | 5.64 | 3.19 |
| p2 | HRV | 7.18 | 8.02 | 6.35 | 7.54 |
| p3 | neg | 0 | 0 | 0 | 0 |
| p4 | HRV | 2.70 | 0 | 0 | 0 |
| p5 | FluA | 7.53 | 6.00 | 6.52 | 5.48 |
| p6 | neg | 0 | 0 | 0 | 0 |
| p7 | neg | 0 | 0 | 0 | 0 |
| p8 | COV | 0 | 0 | 3.99 | 3.29 |
| p9 | COV | 0 | 0 | 0 | 2.70 |
| p10 | COV | 2.70 | 0 | 0 | 0 |
| p11 | neg | 0 | 0 | 0 | 0 |
| p12 | COV | 3.40 | 2.70 | 4.84 | 0 |
| p13 | COV | 0 | 2.70 | 0 | 4.71 |
| p14 | COV | 2.70 | 2.70 | 0 | 0 |
| p15 | RSV | 5.34 | 0 | 3.26 | 0 |

PIV = parainfluenza virus; HRV = human rhinovirus; FluA = influenza virus A; COV = coronavirus; RSV = respiratory syncytial virus

### Performance characteristics of foam versus flocked swabs for measurement of nasal viral load

We first compared the yield of samples collected using foam versus flocked swabs within the same nostril. All study participants provided paired specimens from both nostrils to allow for direct comparison. The agreement between samples collected by foam and flocked swabs was generally high, particularly with high viral load samples, with no evidence of higher yield with one method versus the other **(Fig 1a)**.

**Figure 1:**
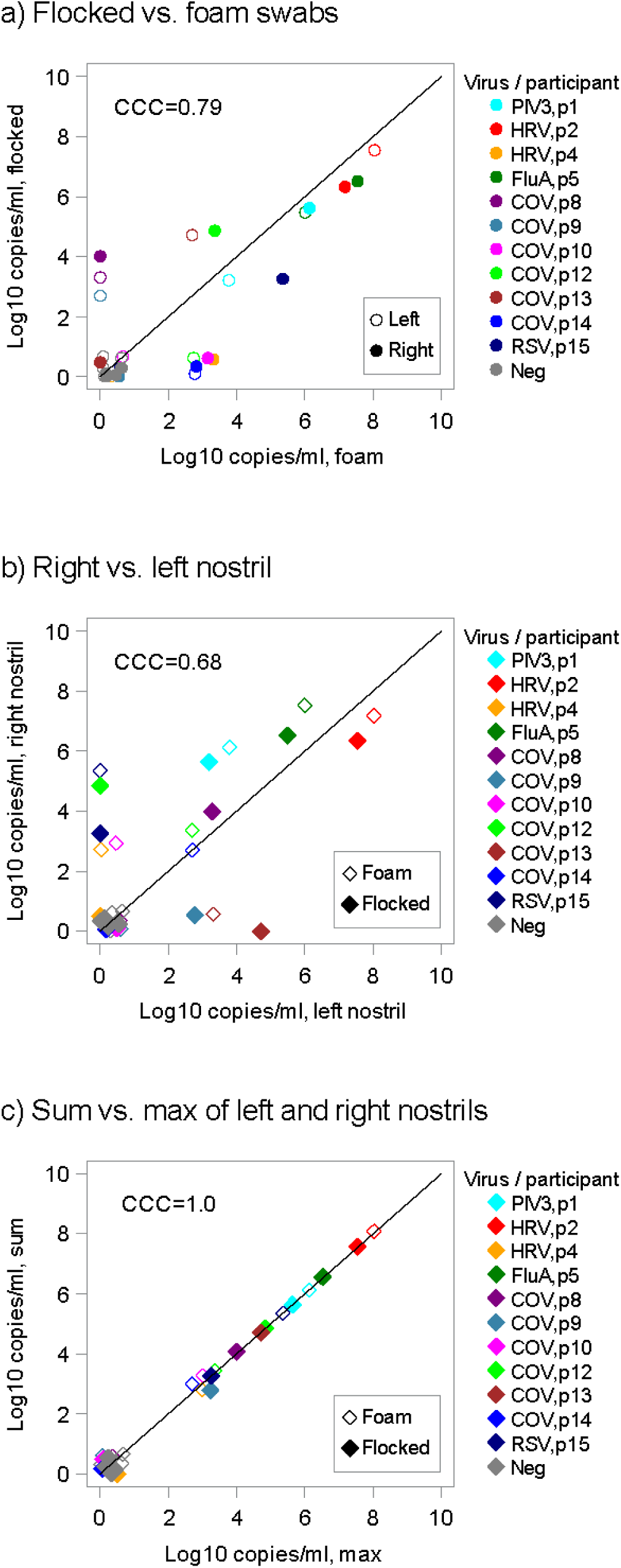
Comparison of viral loads between self-collected foam and flocked swabs. **(a)** Viral loads from the same nostril using flocked and foam swabs are concordant, particularly at higher viral loads. **(b)** Differential viral loads with the same swab type, observed between nostrils, show moderate concordance. **(c)** Viral load from the highest nostril strongly agrees with the sum of the two nostrils suggesting that a majority of sampled virus comes from one side. Overlapping data points have been jittered to allow viewing of all data points. CCC = concordance correlation coefficient; CoV = coronavirus; FluA = Influenza A; HRV = human rhinovirus; PIV3 = parainfluenza virus 3; RSV = respiratory syncytial virus.

### Focality of respiratory virus shedding in nasal passages

In the same dataset, we compared swab samples obtained with the same swab type from separate nostrils with a total of 15 paired samples. The values for these viral loads were notably higher in one nostril than the other and were less in agreement **(Fig 1b)**. Moreover, the value from the highest nostril strongly agreed with the sum of the two nostrils suggesting that a majority of sampled virus comes from one side **(Fig 1c)** and that sampling the other side underestimates viral load. Therefore, bilateral sampling is likely required for optimal yield and accurate quantitation.

### Comfort and ease of self-collected foam swabs compared to flocked swabs

There was a trend suggesting that participants found foam swabs more comfortable (9/15 participants agreed or strongly agreed that the foam swabs were comfortable to use, whereas 4/15 participants agreed or strongly agreed that flocked swabs were comfortable) although this did not reach statistical significance (p=0.13). Foam swabs were also reported to be easy to collect (14/15 participants agreed or strongly agreed for foam swabs vs 11/15 for flocked swabs; p=0.25).

Almost all participants (14/15) would consider participating in future research using foam swabs, but only 10/15 if flocked swabs are used (p=0.13).

### Ease, comfort and high compliance associated with longitudinal nasal sampling during an upper respiratory virus infection

We next enrolled a cohort of 9 otherwise healthy, adult study participants who self-sampled their nasal passage serially for 14 days, starting within 3 days of upper respiratory symptoms. One participant contributed serial samples twice. Overall compliance was high: median number of sample days was 14 (range 11-19 days). After completion of the sample collection period, 70% of participants agreed or strongly agreed that the foam swab was comfortable, 90% agreed or strongly agreed that the foam swab was easy, and 80% agreed or strongly agreed that they would participate in future research with foam swabs. Additionally, 80% of participants agreed or strongly agreed that the swab collection instructions were easy to follow, and 90% agreed or strongly agreed that the collection kit return process was easy. Serial home-based testing appears to be a well-accepted methodology.

### Steady-state nasal passage viral load kinetics during respiratory virus infections

In the longitudinal sampling portion of our study, we were able to detect 14 viruses including seven human rhinovirus (HRV), two coronavirus (CoV), one bocavirus (BoV), two adenovirus (ADV), one human metapneumovirus (MPV) and one respiratory syncytial virus (RSV) cases. There were four instances of viral co-infection, though in each case a dominant virus was evident based on greater duration of shedding and higher viral load **(Fig 2a)**.

**Figure 2:**
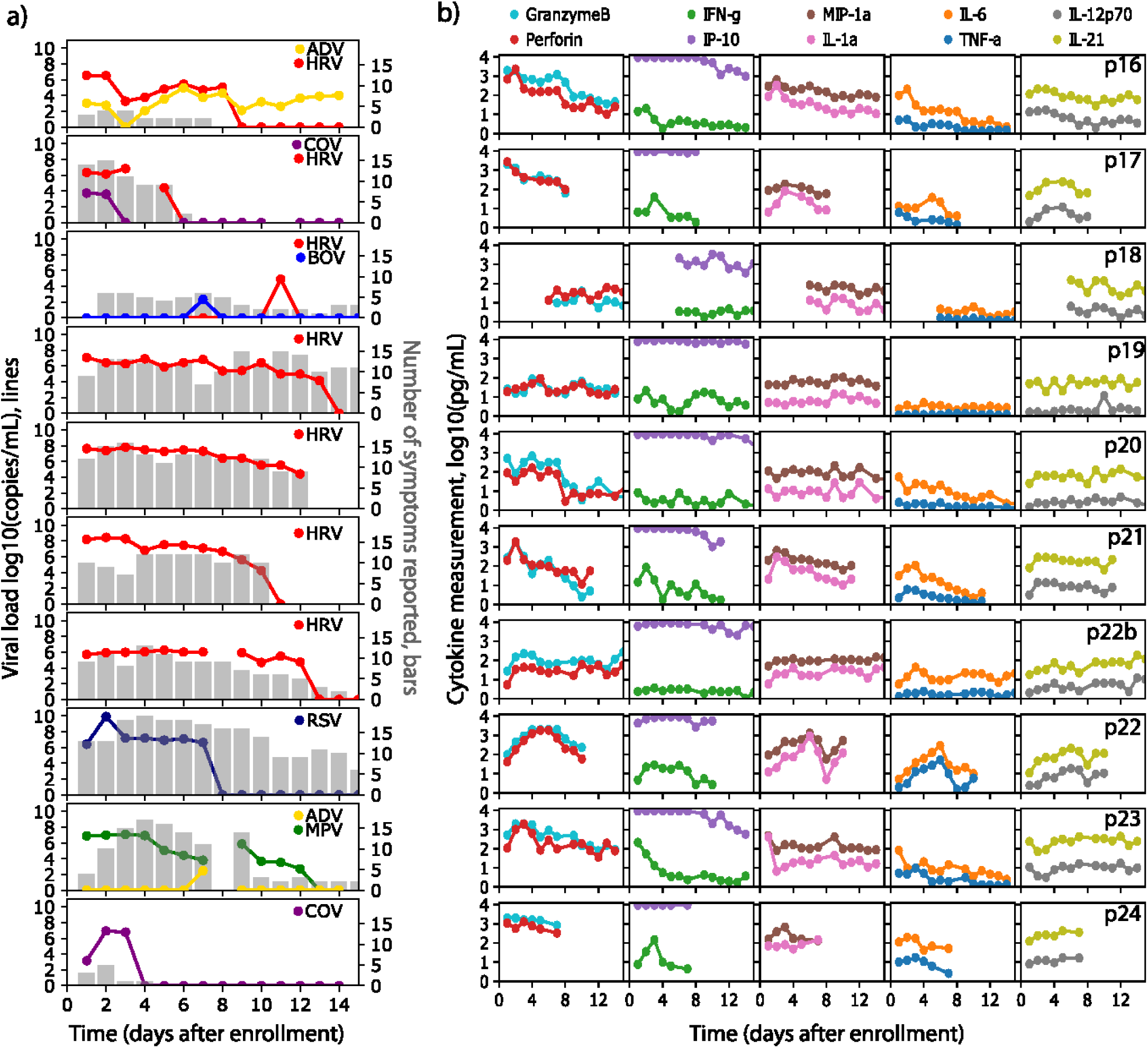
Viral load, symptoms and cytokine levels in serial sampling in both nostrils. Each row represents a participant. **(a)** Viral load (lines) and quantity of symptoms (bars) are shown on left and often tracked with each other longitudinally. Serial sampling in both nostrils with foam swabs reveals a steady state for HRV, RSV and HMPV viral loads prior to rapid elimination. **(b)** Levels for each cytokine (granzyme B, perforin, IFN*γ*, IP-10, MIP-1α, IL-1α, IL-6, TNF-α, IL-20p70, IL-21) are shown on the right. Paired cytokines show concordant expansion and clearance phases. HRV = human rhinovirus; RSV = respiratory syncytial virus; MPV = metapneumovirus; ADV = adenovirus; CoV = coronavirus, BoV = bocavirus.

Duration of shedding was heterogeneous. In 5 cases, HRV shedding lasted more than a week with one instance of 5-day shedding and one short single-day blip. RSV and MPV episodes were both prolonged. ADV, BoV and CoV shedding was short-lived, though in one case low-level ADV shedding was evident throughout the sampling period **(Fig 2a)**.

During most extended periods of HRV, RSV and HMPV shedding, viral loads were remarkably stable from sample to sample **(Fig 2a)**. For HRV, a generalized pattern of viral load steady state or slight gradual decline, followed by rapid elimination was noted. The single case of RSV had a similar profile but with an initial high viral load peak and shorter duration of shedding. The single case of MPV had a more protracted decline with a single re-expansion phase. These transiently observed periods of steady state viral loads are highly unlikely to occur by chance if true viral loads fluctuated or exhibited stochastic noise. Thus, the sampling method appears highly reliable. These data also suggest a brief period of equilibrium between the virus and local immune system before viral elimination.

### Viral load kinetics as a predictor of respiratory virus symptoms

In general, the level of symptoms appeared to track with detectable virus, particularly for COV, HRV and MPV. For the single case of RSV, a high number of symptoms persisted beyond viral elimination **(Fig 2a)**. For all HRV infections of greater than one day, duration of shedding correlated strongly with duration of symptoms (r=0.87). In these HRV infected individuals, symptoms subsided immediately before, concurrent with or soon after viral elimination. Low viral load infections lasting only a day were associated with a smaller number of symptoms than more prolonged higher viral load episodes **(Fig 2a)**.

### Stable and surging nasal cytokine levels during respiratory virus infection

We next followed the levels of 20 different cytokines during infection measured from the same specimens from which the viral load was measured. For several cytokines particularly those in the Th2, Th17 and non-defined pathways (IL-2, IL-4, IL-5, IL-10, IL-13, IL-17A and eotaxin), there was notable stability within and between study participants, independent of viral shedding **(Fig 2b, Supp Fig 2b)**. Interferon α2a also was relatively invariant across and within persons. This result demonstrates consistency in swabbing technique and yield for molecules that are not impacted by the presence of viral infection, and again validates the precision of our approach.

Other molecules, particularly those associated with cytotoxic T cell responses (granzyme B, perforin, TNFα and IFN*γ*) and macrophage responses (MIP-1α, IL-1α, IL-6, IL-18) showed monotonic expansion or clearance in response to most infections, with particularly dynamic shifts during the examples of RSV, HPMV and one instance of HRV (p21) with the highest initial viral load **(Fig 2b, Supp Fig 2b)**. Other cytokines such as Il-12p70, IL-21, IL-5 and IL-18 were dynamic in participants had monotonic expansion and clearance in response to some but not all infections.

### Cytokines correlations according to cellular origin

We next examined the six participants with HRV infection for any correlated cytokine patterns to infer cellular origin. Examples of high positive correlations were noted among analytes associated with a cytotoxic T cell response (granzyme B, perforin, TNFα, and IL-6), among several macrophage or epithelial cell derived cytokines (MIP-1α, IL1α, IL-6, IL-12p70, IL-21), and among a cluster associated with Th2 responses (IL-5, IL-10, IL-17). The Th2 associated cytokines (IL-5, IL-10, IL-17) also correlated with many of the cytolytic T cell and macrophage associated cytokines **(Fig 3a)**. HRV viral load was only moderately correlated with a number of parameters, particularly granzyme B, perforin, and IP10 which is a protein induced by IFN*γ*. This result suggests that HRV may not induce an intense local molecular immune response in a dose-dependent fashion. Very similar results occurred with inclusion of all samples from all participants in the cohort **(Supp Fig 3a)**.

**Figure 3:**
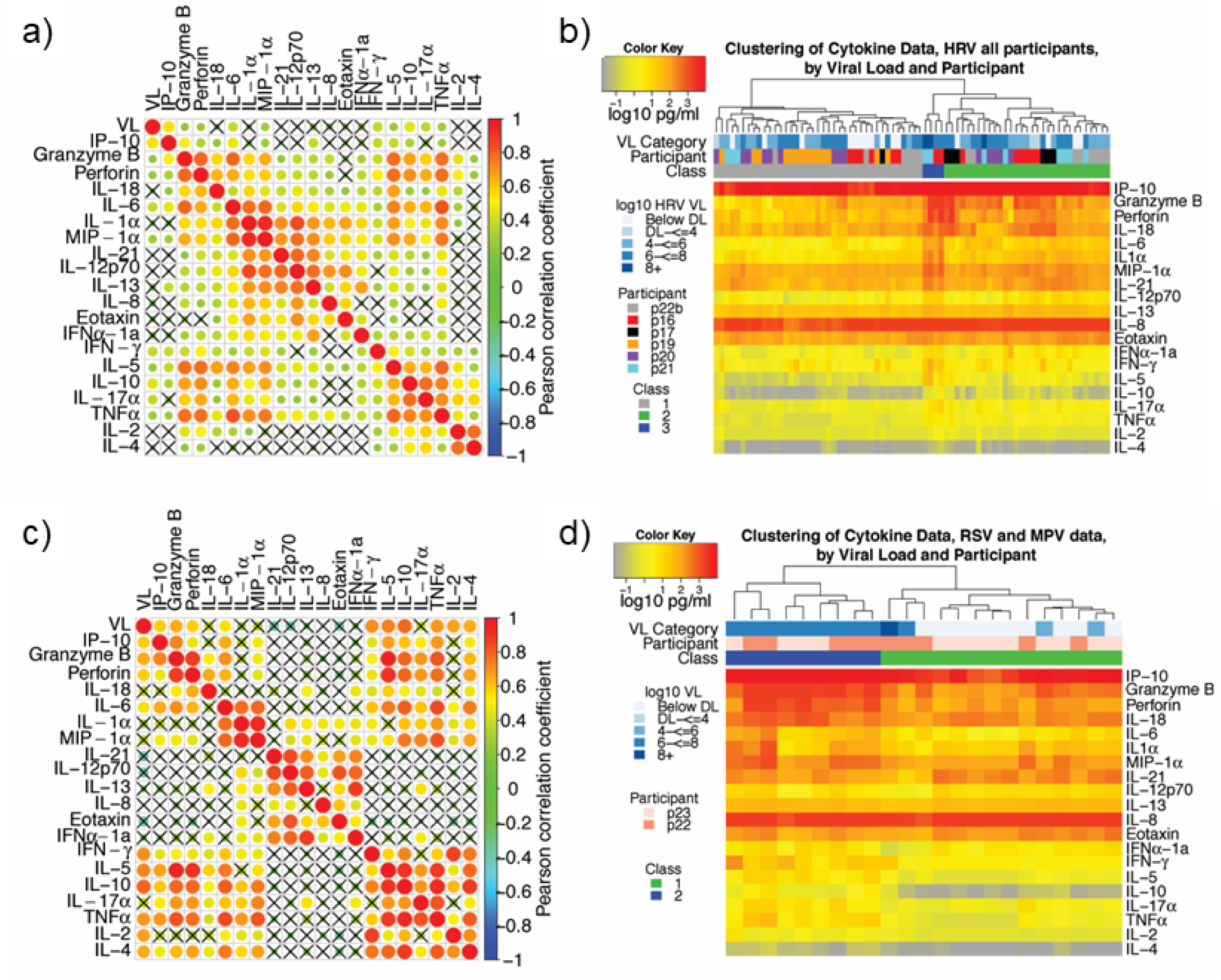
Cytokines correlate according to cellular origin during respiratory virus infection, while samples cluster according to level of inflammation. **(a, b)** Data from participants p16, p17, p18, p19, p20, p21 and p22b who have HRV infection. **(c, d)** Data from participants p22 and p23 who have RSV and MPV respectively. **(a, c)** Correlation plots with strong correlation according to cell type origin. X indicates a non-significant correlation. Color intensity and the size of the dot are proportional to the Pearson correlation coefficient. For both datasets, strong positive correlations are noted within cytokines linked with cytolytic T-cell responses; macrophage responses; and T_H_2 responses. **(b, d)** Linkage clustering analysis of samples (columns) demonstrates classes of samples based on the concentration of inflammatory cytokines. **(b)** For HRV infections, a minority of samples (blue class) from 2 participants and with the highest levels granzyme B, perforin, IL-6, IL-1α, MIP-1α and IFN*γ* all had high viral loads. All six participants had samples in the least inflammatory class (grey) and five participants had samples in the moderate inflammatory class (green). **(d)** For RSV and MPV, inflammatory (blue) and non-inflammatory (green) sample clusters are evident. The inflammatory class of samples is highly associated with the highest viral loads. VL = viral load; DL = detection limit.

We next examined cytokine correlations in the 2 persons infected with more inflammatory respiratory viruses: RSV and MPV. We noted similar correlative trends in this data as with HRV. Correlations among related pairs such as granzyme B / perforin, IL-12p70 / IL-21, IL-1 / MIP-1alpha, IL-5 / IL-18, TNF-alpha / IL-6, IL-10 / IL-17a and IL-2 / IL-4 were higher for RSV / MPV than for HRV **(Fig 3b)**. For these cytokine pairs, temporal kinetics were often strikingly similar suggesting an equivalent cellular source **(Fig 2b, Supp Fig 2b)**. There was also an overall lack of correlation between cytokines associated T cells responses and those associated with epithelial cells and macrophages. Viral load correlated with many cytokines of T cell origin **(Fig 3b)**, suggesting that RSV and MPV may induce inflammation is a dose-dependent fashion.

### Sample clustering according to degree of inflammation

We next sorted all HRV samples using linkage clustering analysis. This approach demonstrated three classes of samples that were distinguished by the levels of many of the T-cell and macrophage-associated molecular immune factors **(Fig 3b)**. The minority of samples (blue class) with the highest levels of granzyme B, perforin, IL-6, IL-1α, MIP-1α and IFN*γ* all had high viral loads. These samples were from two participants. All six participants had some samples in the least inflammatory class (grey) and 5 participants had samples in the moderate inflammatory class (green). These data indicate that the inflammatory immune milieu in the HRV-infected nasal passage is dynamic over time, but tilts toward higher inflammation with higher viral loads. Very similar results were observed when all samples were analyzed though only two classes of samples were distinguished **(Supp Fig 3b)**.

We next sorted the RSV and MPV samples using linkage clustering and could not identify the optimal number of clusters. We selected two clusters which were differentiated according to concentrations of most cytokines, again including granzyme B, perforin, IL-6, IL-1α, MIP-1α and IFN*γ*. In the case of these viruses, the more inflammatory cytokine cluster clearly associated with high viral loads for both RSV and MPV **(Fig 3d)**.

### Mathematical modeling

We performed mathematical modeling separately on data from the participant infected with RSV to examine whether complex immune and viral data from our samples could be coupled mechanistically. We first developed the ordinary differential equation model in equation (1) to link RSV viral load and early and late immune responses and evaluated which cytokines may track those responses. For the early immune response, we found that only the log_10_ of the concentration change of IFN-γand IP-10 was positively correlated to the viral load during the first 5 days after enrollment **(Supp Fig 4a-b)**, so we evaluated models for only these two cytokines to track the RSV-early immune response. For the late immune responses, we evaluated the model for fit to all observed cytokines **(Supp Fig 4c)**. An equivalent approach was carried forward to model MPV.

**Figure 4:**
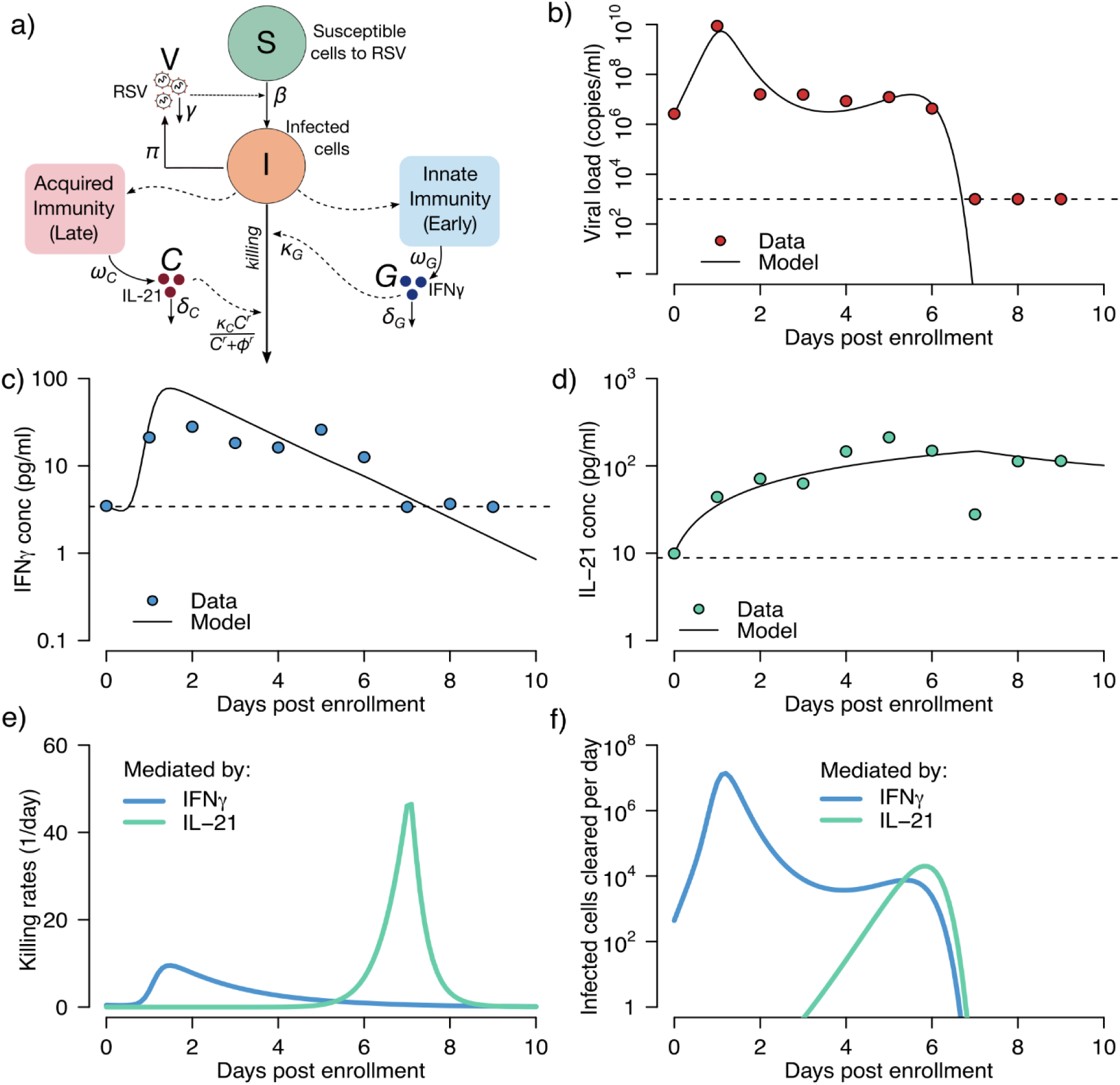
Mathematical modeling of a single participant’s RSV kinetics and the early and late immune responses tracked by IFN-γand IL-21. **(a)** Schematic representation of the model. *S* represents cells susceptible to RSV; *I*, RSV-infected cells; *V*, RSV virions; *G*, IFN-γconcentration and *C*, IL-21 concentration. Best fit models to **(b)** viral load, **(c)** IFN-γand **(d)** IL-21 measurements using a nonlinear least-squares approach. Circles represent the data, and black-solid lines the best model predictions. Models fit better to these cytokines than all others charted in **Fig 2** and **Sup fig 2. (e)** Model estimates of the killing rate per cell of infected cells mediated by IFN-γand IL-21, calculated as *κ* _*G*_ *G* and 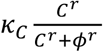, respectively. **(f)** Total number of infected cell deaths mediated by IFN-γand IL-21, computed as *κ* _*G*_ *GI* and 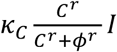, respectively. In **(e)** and **(f)** blue and green lines represent model predictions of the effects mediated by IFN-γand IL-21, respectively.

In **Fig 4a**, we show our resulting model schematic. Differential equations capture the rate of change of susceptible cells, infected cells, viral load **(Fig 4b)** and two cytokines **(Fig 4c-d)**. The best fit was achieved with a model assuming IFNγ concentration dependent killing during the first early stages of infection, and IL-21 mediated elimination of infected cells with a mechanism saturating the amount of possible killing above a certain level of IL-21.

The model suggests that for RSV, an early surge in IFNγ leads to a slight rise in per cell killing rate of infected cells **(Fig 4e)** leading to a mass elimination of infected cells at a rate of 10 million cells per day and a decrease in viral load by a factor of 100-1000. However, this response does not clear the virus. A steady state viral load persists for 4 days until an IL-21 mediated response appears. This response kills far more rapidly, but not as intensely as the IFNγ response. Together, these responses remove the remaining infected cells by day 7 after onset of symptoms **(Fig 4f)**. Model fitting using data in which IP10 provides early clearance rather than IFNγ results in worse fit to the data **(Supp Fig 4c, 4d)**. For late results, IL-21 allowed the best fit and other cytokines was less successful. This suggests that these two molecules may play a major role in RSV control in vivo but does not rule out the effects of other cytokines and effector molecules in limiting infection.

We next fit the same model to the data from participant infected with MPV and found that the model is able to recapitulate viral load, IFNγ and IL-21, projects similar killing patterns during the early and late immune responses to the RSV model **(Supp Fig 5)**.

## Discussion

Here we demonstrate that home self-sampling with nasal foam swabs is well-tolerated and provides reliable results for monitoring viral load as well as the molecular immune response to respiratory virus infection. These results have enormous practical implications. Self-collection at home is safe, non-invasive and easily learned, allowing a reliable method for diagnosis as well as therapeutic monitoring. Because our kits could easily be used at home or in a drive through testing environment, they provide an avenue to eliminate contact between an infected and contagious person, and health care providers. They could also be used in the hospital or clinic setting, thereby saving personnel time and personal protective equipment. The use of comfortable, safe and affordable foam swabs also highlights the possibility of scaling this approach to pediatric, adult, elderly and immunocompromised populations. For the current SARS-CoV-2 pandemic, and future deadly respiratory virus epidemics, home self-swabbing will be a vital tool.

The simplicity of the sampling approach also facilitates large scale research studies of viral pathogenesis and transmission dynamics in which participants self-sample for months. Our inability to stop the spread of the COVID-19 epidemic in the United States has demonstrated a poor overall understanding of cryptic transmission patterns of respiratory viruses. Because our approach is safe, well accepted, and easy to implement, longitudinal sampling studies within families, workplaces and at large conferences are highly feasible.

We have previously demonstrated increased sensitivity of self-collected foam nasal swabs compared to nasal washes in immunocompetent adults with respiratory viral infections^24^. Additional anatomical sites have also been considered for increasing yield, and current FDA recommendations suggest use of both a mid-turbinate nasal swab and an oropharyngeal swab to maximize yield in the absence of nasopharyngeal swabbing^23^. Our prior data demonstrate that self-collected throat swabs in addition to self-collected foam nasal swabs do not increase yield significantly for respiratory viruses^26^, suggesting that additional oral swabbing may not be needed, especially in the setting of swab shortages. Self-collected foam swabs have been used for longitudinal studies in solid organ transplant recipients^25^, with good compliance and participants reporting no issues with swab discomfort. The specific swab used in these prior studies and our present study were custom designed to limit discomfort while maintaining adequate sensitivity; we have demonstrated stability with these swabs with and without transport media after storage at room temperature for 7 days^24^, making them ideal for home self-testing followed by shipment directly to a testing lab. Furthermore, SARS-CoV-2 has been shown to be highly stable on surfaces^27^, making home foam swabbing a feasible and attractive option for this pathogen.

We also demonstrate an ability to accurately sample local cytokines which are present at picogram levels, again using the same foam swabs from which viral measurements were made. The combination of precise virologic and immunologic readouts of local infection is highly relevant for developing clinical severity scores and biomarkers. While studies are beginning to show that viral load may be predictive of COVID-19 severity^10^, it is equally plausible that the intensity and phenotype of the early local cellular immune response plays a causal role in limiting the extent of infection^28^. By following the molecular immune response closely with daily sampling intervals, we also provide adequate data for mathematical models that can link specific arms of the cellular immune response to pathogen control in real time^22^, a goal that has been difficult to attain for a majority of viral infections in humans.

Our study demonstrates several novel features of respiratory virus kinetics. RSV infection achieves a brief, extremely high, viral load, followed by a steady state and a final rapid phase of elimination. HRV also has a remarkably stable viral load in most participants before being rapidly eliminated. During a majority of our observed episodes, viral shedding is strongly correlated with symptoms. As viral load decreases, symptoms tend to dissipate.

Certain molecular immune responses are constitutively expressed, and vary little between and within participants, particularly those associated with Th2 mechanisms that are unlikely to play a role in elimination of virally infected cells. On the other hand, small molecules associated specifically with tissue-resident T cell responses such as granzyme B, perforin and IFNγ, and macrophages such as IL-6 and IL-1 expand and contract during the course of viral shedding, particularly with more severe infections such as RSV and HPMV. Our technique therefore overcomes a fundamental limitation of human immunological studies, which is the inability to sample over temporally granular time intervals at the mucosal site of viral replication.

Further validation of our technique is demonstrated with mathematical modeling that links expression of certain cytokines with early and late elimination of virus. For RSV and MPV, we demonstrate that an early surge in IFNγ is coupled with elimination of a massive number of infected cells but is insufficient for complete containment of infection, which is achieved several days later concurrent with slower expansion IL-21. Notably, IL-21 has previously been identified as required for RSV elimination in murine models^29-31^ In our model, it induces an extremely high death rate of infected cells once above a certain concentration. Larger scale studies may be able to link surges in different cytokines with different respiratory viruses, including SARS CoV-2, and to differentiate severity using these techniques. Of particular interest is combining information on levels of *local* cytokine levels with viral load at presentation, along with patient metadata, to predict infection severity.

There are important limitations to our study. Correlations between foam and flocked swabs were weaker at low viral loads. However, stochastic variation in low viral load samples is inherent to quantitation of viruses which replicate in mucosa. Additional variables such as storage temperature may have contributed to viral quantification variability. Our samples size for longitudinal episodes is relatively low, particularly when considered on a per virus basis. A greater number of participants will be required to definitively differentiate kinetics patterns of different respiratory viruses, as well as the cytokine profiles associated with their containment. Selection of cytokines as incomplete and may have missed critical responders to viral infection. Our mathematical models dramatically oversimplify the coordinated immune response against the virus but do generate testable hypotheses that IFNγ and IL-21 are viral for early and late containment of infection.

In summary, we establish a foam swab-based sampling method that is optimal for patient self-testing, both at home and in the clinical setting, permits serial therapeutic monitoring, and is suitable for tracking the natural virologic and immunologic course of respiratory virus infections. We recommend that this method be adapted to future clinical and research applications, including for the study of SARS-CoV-2.

## Methods

### Protocol

The study was approved by the Institutional Review Board at Fred Hutchinson Cancer Research Center.

#### Flocked vs foam swab study

Participants with symptoms of an acute respiratory illness, defined as the presence of respiratory symptoms **(Supp Table 1)** for less than 3 days, were enrolled in the study. Each participant completed 2 sample collections, each separated by one hour. At each time point, the participant collected either a) two self-collected Copan flocked swabs (#23-600-966), one from each nostril or b) two self-collected Puritan foam swabs (Puritan Medical Red #25-1805-SC 2), one from each nostril. The foam swab was designed in a mushroom shape to maximize swabbing from the nostril wall **(Supp Fig 1)** and has been used in previous studies in HCT and lung transplant recipients^24,25^. Foam and flocked swabs were self-collected following instruction by trained study personnel. Participants used a saline spray bottle with a nozzle to dispense 5 sprays into one nostril. The participant then placed the swab into the moistened nostril and rotated the swabs and blew for about 5 seconds or 5 rotations. Following sample collection, participants were asked to complete a brief survey to assess the tolerability and acceptability of the various testing methods.

Immediately following collection, each nasal swab was placed in a conical vial containing 1000ul of cytokine preservative buffer consisting of phosphate buffered saline (PBS) with 10% Igepal, 1% protease inhibitor cocktail (EMD Millipore: 539131-1VL), and 0.25% bovine serum albumin (BSA; Sigma A7906-100G). All swabs from the right nostril were stored at -20°C; all swabs from the left nostril were stored at 4°C. All samples were stored for 1 week prior to processing. Swab collection order (flocked vs foam) was randomized using an online randomization tool (www.randomizer.org). To compare the number of participants who agreed or strongly agreed with statements regarding comfort, ease of use, and participation in future research for foam versus flocked swabs, we used McNemar’s test with exact p-values.

#### Longitudinal sampling study

Participants with symptoms of an acute respiratory illness, defined as presence of respiratory symptoms **(Supp Table 1)** for less than 3 days, were enrolled in the study. Each participant collected two Puritan foam nasal swabs, one from each nostril, per day for 14 days after enrollment or until symptoms resolved, whichever was longer. Participants completed a daily electronic symptom survey, in which participants were asked to record the presence and severity of symptoms in specific categories: nasal, eyes, ears, throat, chest, gastrointestinal, general, sleep and sensory changes **(Supp Table 1)**. Following completion of the 14-day sample collection, participants were asked to complete a brief survey to assess the tolerability and acceptability of the testing methods.

Immediately following collection, each nasal swab was placed in a conical vial containing 1000ul of cytokine preservative buffer consisting of 0.1% Tween 20, 1% protease inhibitor (EMD Millipore: 539131-1VL), 1% BSA (Sigma A7906-100G), 1X ProClin300 (at 1:2000, diluted with PBS). Participants were instructed to store collected swabs in the participant’s home refrigerator. Participants then transported collected samples to the lab in insulated bags containing ice packs within one week of sample collection. Nasal swabs were processed within one week of sample collection. Collections from each nostril were combined for the final analyses.

### Lab methods

#### Sample processing

Each conical vial containing a swab was vortexed and 500ul of buffer was removed and stored at -80°C for PCR analysis. The swab was then removed from the conical vial and placed in a pre-chilled 0.45um SPIN-X filter and the handle of the swab was removed. The buffer remaining in the conical vial was then transferred to the SPIN-X filter containing the swab. The SPIN-X filter was then spun at 13000xg for 15 minutes at 4°C with no brake. 300ul of fresh cytokine preservative buffer was then added to the SPIN-X filter which was then incubated on wet ice for 5 minutes then spun again at 13000xg for 30 minutes at 4°C with no brake. The swab and filter were discarded, and the filtered buffer was then aliquoted in 100ul increments and stored at -80°C until further testing.

#### Viral testing

Nasal swab specimens were tested using a multiplex PCR testing for 11 respiratory viruses [adenovirus A-F, human rhinovirus (HRV), influenza A and B, parainfluenza viruses (PIV) 1-4, human coronavirus (CoV), bocavirus (BoV), respiratory syncytial virus (RSV) and human metapneumovirus (MPV)] as previously described^32^.

#### Cytokine testing

Cytokine levels were quantified in nasal specimens using the electrochemiluminescence-based Mesoscale Discovery (MSD) platform. For the longitudinal sampling study, the following panels were used: U-PLEX Biomarker Group 1 (Eotaxin, IFN-α2a, IL-1, IL-8, IL-12p70, IL-13, IL-18, IL-21, IP-10, MIP-1α), U-PLEX Custom Biomarker (IFN-γ, IL-2, IL-4, IL-5, IL-6, IL-10, IL-17A, TNF-α), R-PLEX Granzyme B, and R-PLEX Perforin. Preparation of analyte detection plates was done following the manufacturer’s instructions (Meso Scale Diagnostics). A series of 8 concentrations of biomarkers standards and the test samples were added in duplicates to the wells. The plates were incubated shaking for 1 hour. In parallel to plate incubation, the plate-respective SULFO-TAG labeled detection antibodies were combined. The plates were washed, and the respective detection antibody mixture was added to each well. The plates were incubated shaking for 1 hour. Plates were washed, then 2X Read buffer was added to each well. The plates were read on the MSD Plate reader (MESO QuickPlex SQ 120). Protein concentrations were determined using the MSD Discovery Workbench 4.0 analysis software. The light intensities from samples were interpolated using a four-parameter logistic fit to a standard curve of electrochemiluminescence generated from the known concentrations of the standards. The lower limit of detection for each marker can be found on the manufacturer’s website: https://www.mesoscale.com/∼/media/files/handouts/assaylist.pdf.

### Statistical analysis

For the foam versus flocked swab study, PCR results that were positive on the qualitative assay but below the limit of detection were imputed as 500 copies per ml, using the limit of detection divided by two. PCR results were log10-transformed and negative results were assigned a value of 0. The concordance correlation coefficient (CCC) was used to measure agreement of quantitative results between paired samples (foam versus flocked swabs, left versus right nostril samples, sum versus maximum value from left and right nostrils)^33^. For the longitudinal sampling study, cytokine results that were below the fitted curve range were assigned the value of the lower limit of detection divided by two and results that were above the fitted curve range were assigned the value of the upper limit of detection.

Results were log10-transformed for analysis. Symptoms are represented as the total number of symptoms present for each day, out of a total of 26 **(Supp Table 1)**. SAS, version 9.4 (SAS Institute, Cary, North Carolina) and Stata, version 16.1 (StataCorp, College Station, Texas) were used for analysis.

### Cytokine clustering

To check whether the samples could be classified into groups with similar cytokine concentrations, we performed a cluster analysis of the samples where each sample is an array of the 20 measured cytokine concentrations. First, we checked for cluster tendency of the samples using Hopkin statistic (H)^34,35^. H can have values between 0 and 1, where values close to 1 indicate that the samples are highly clustered and values close to 0.5 indicate random samples. When calculated H (get_clust_tendency function in R3) was greater than 0.5, we did a linkage hierarchical clustering with Euclidean distances of the samples^36^.

### Mathematical modeling

*Model assumptions:* To understand how the immune system drives respiratory virus dynamics we used an acute viral infection model that distinguishes between early and late responses to RSV. In this model, susceptible cells (*S*) are infected at rate *βVS* by free RSV virus (*V*). The impact of host immunity is tracked by modeling two cytokines that are plausible surrogates for those responses. We assumed RSV-infected cells (*I*) are cleared by: (1) an innate response with rate *κ*_*G*_ *G* mediated by an innate immune response tracked by an initial cytokine (*G*); and (2) an acquired response with rate 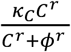 mediated by an acquired immune response tracked by a second cytokine (*C*). The Hill coefficient *r*parameterizes the nonlinearity of the response and allows for rapid saturation of the killing. In the model, *G* is secreted proportionally to the number of infected cells with rate *ω*_*G*_*I* and cleared with rate *δ*_*G*_*G*. *C* is secreted in a non-linear fashion with density dependent rate 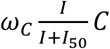 and cleared with rate *δ*_*C*_ *C*. Finally, free virus is produced at a rate *π* and cleared with rate *γ*. The model is expressed as a schematic **(Fig 4a)** and here as a system of ordinary differential equations:

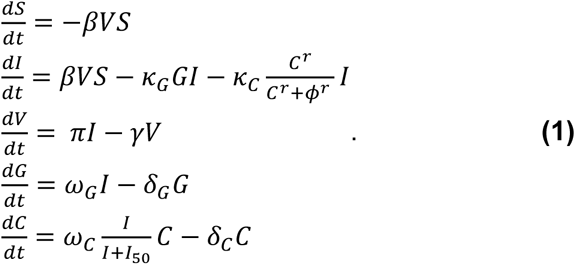

*Selection of surrogate cytokines:* To select the initial cytokine to model the surrogate for early immune response (*G*) we performed Pearson’s correlation tests between the RSV viral load from day 1 until day 5 post-enrollment and the log_10_ of the concentration change of each cytokine until day 5 post-enrollment. We modeled equation (1) only for the cytokines with positive correlation that were statistically significant. Then for each cytokine obtained for variable *G* we tried model fitting to all cytokines for variable *C* individually. We selected the surrogate for the acquire immune response to RSV (*C*) the cytokine that gave a lower sum of squares error in the model fitting.

#### Model fitting: Model fitting

We performed fitting of model in equation (1) to the data assuming *t*=0 as the time of enrollment. We also assumed initial concentrations of *S*(0) =10^7^cells/μL,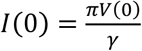 and obtained *V*(0), *G*(0) and *C*(0) from the viral load and cytokine initial concentrations, respectively. We estimated the remaining best parameters using nonlinear least-squares. Implementation used the differential evolution (DEoptim) and the L-BFGS-B (optim) algorithms in R.

#### Model predictionsx

We used equation (1) and best estimates from the best model fits to calculate the absolute number of infected cells killed and the killing rate per cell during early and late immune responses against RSV. The number of eliminated infected cells at any time was calculated by the equations *κ*_*G*_*GI* and 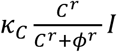 for the early and late responses, respectively. Similarly, we computed the killing rate per cell during early and late response as *κ*_*G*_ *G* and 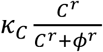.

## Data Availability

The data that support the findings of this study are available from the corresponding author upon reasonable request.

## Acknowledgements

We would like to thank our study participants.

## Author Contributions

AW designed the experiments and wrote the manuscript. EMK performed statistical analysis, SB performed statistical analysis and mathematical modeling. EV performed data analysis. TL enrolled participants and performed experiments. ELC enrolled participants and performed experiments. UP performed the cytokine analysis. JK performed respiratory virus PCR. ERD wrote the manuscript. KRJ designed the respiratory virus PCR. ALG performed the respiratory virus PCR. DBR performed mathematical modeling. EFCO performed mathematical modeling. MB designed the experiments and wrote the manuscript. JTS designed the experiments, designed the mathematical modeling and wrote the manuscript.

## Competing Interests Statement

A.W.: Kyorin (personal fees), Ansun (research support), VB Tech (research support), all outside of the submitted work. A.G.: Abbott Molecular, personal fees, outside of the submitted work.

M.B.: Kyorin (personal fees), Gilead (research support, personal fees), ReViral (personal fees_, Janssen (research support, personal fees), Ansun (research support, personal fees), Moderna (personal fees); Vir Bio (research support, personal fees); GSK (personal fees), Pulmocide (personal fees), VB Tech (research support), Bavarian Nordic (personal fees), DMA (personal fees), Allovir (personal fees), all outside of the submitted work.

**Supplementary Figure 1:**
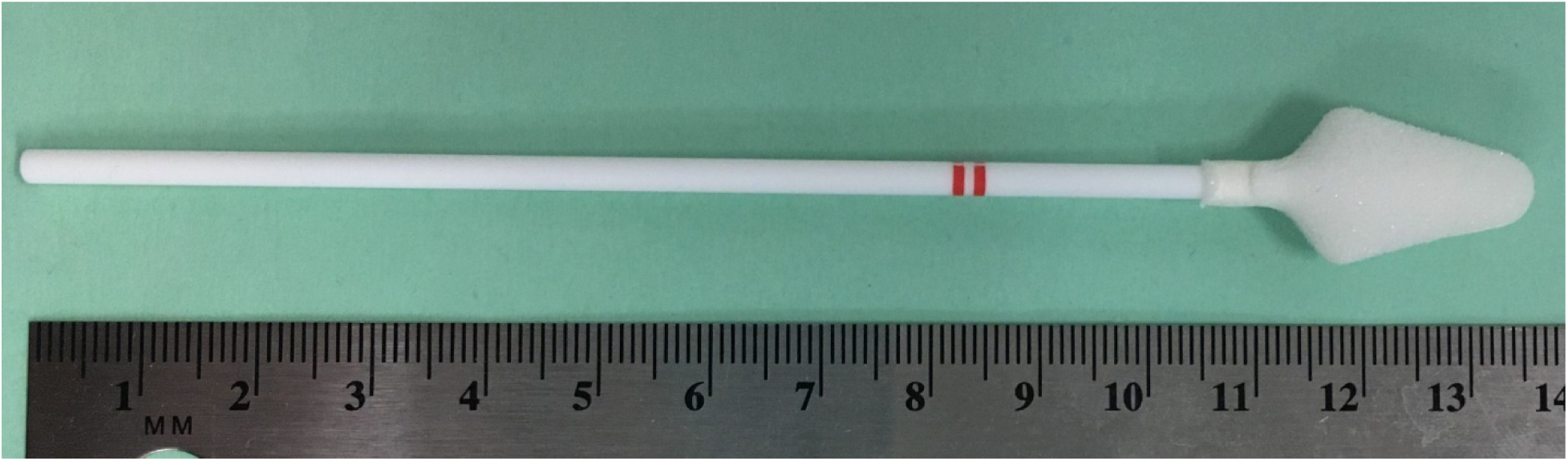
Image of Puritan foam swab (Puritan Medical Red #25-1805-SC 2) used in swab comparison and longitudinal sampling study.

**Supplementary Figure 2:**
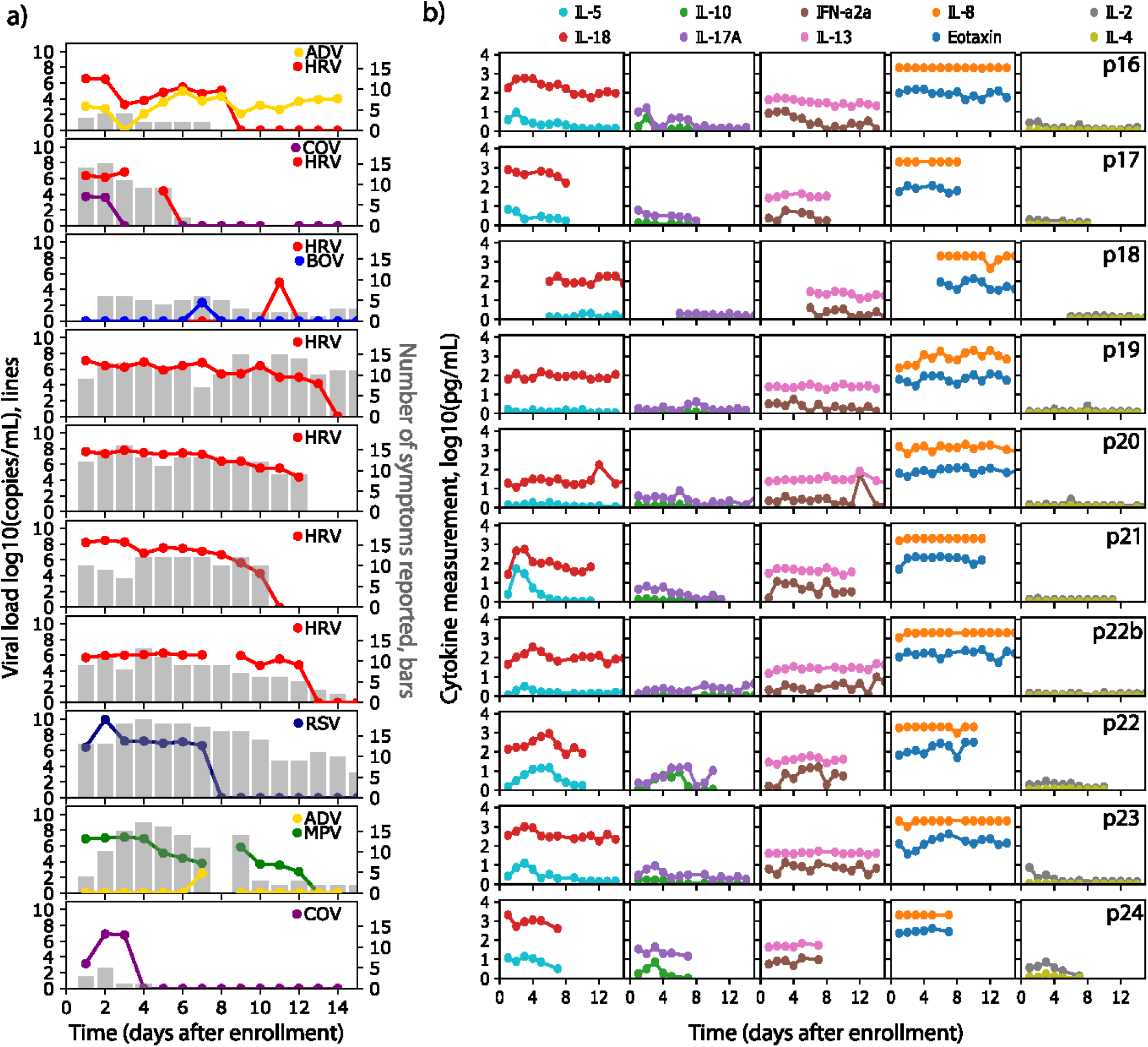
Symptoms, viral load and cytokine levels in serial sampling in both nostrils in all participants for remainder of cytokines evaluated. Each row represents a participant. **(a)** Viral load (lines) and quantity of symptoms (bars) are shown on left and often tracked with each other longitudinally. Serial sampling in both nostrils with foam swabs reveals a steady state for HRV, RSV and HMPV viral loads prior to rapid elimination. **(b)** Levels for each cytokine are shown on the right. Paired cytokines show concordant kinetics. HRV = human rhinovirus; RSV = respiratory syncytial virus; MPV = metapneumovirus; ADV = adenovirus; CoV = coronavirus, BoV = bocavirus.

**Supplementary Figure 3:**
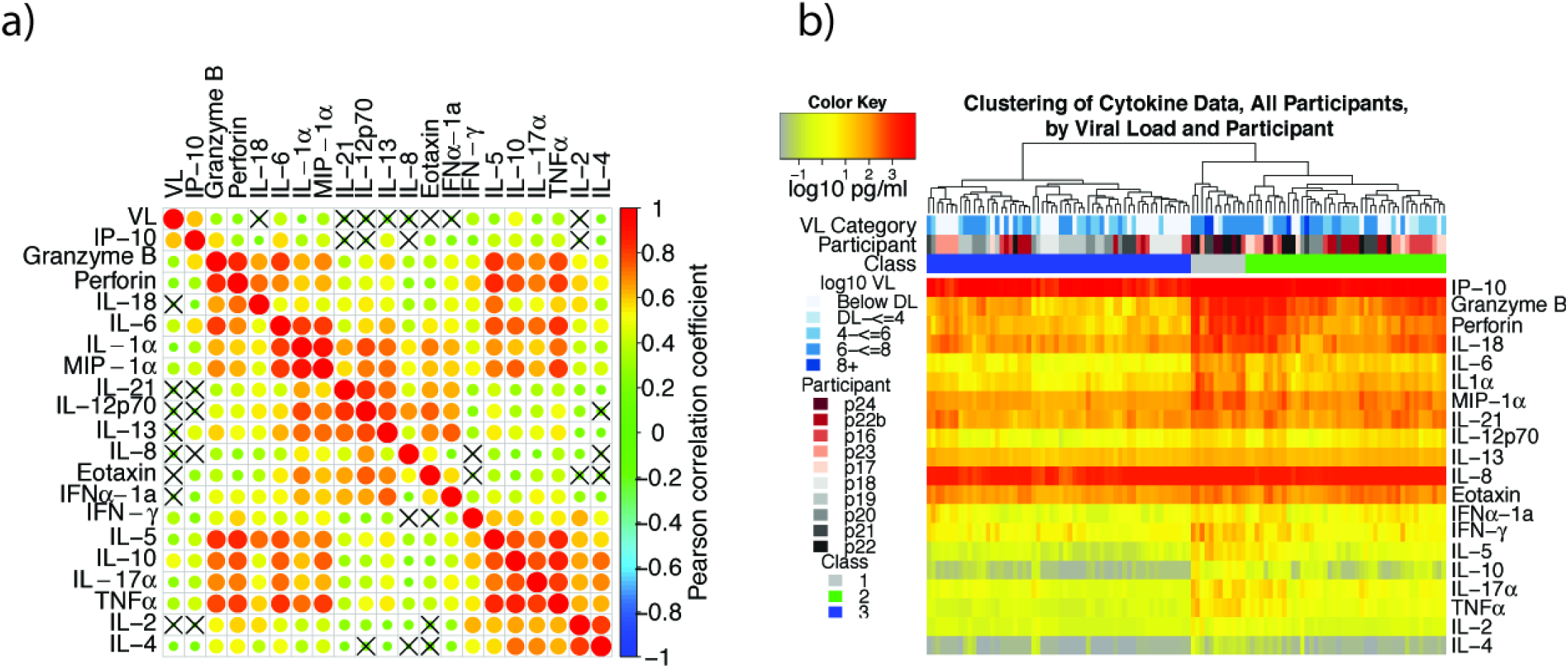
Cytokines correlate according to cellular origin during respiratory virus infection, while samples cluster according to level of inflammation. Data is from all participants. **(a)** Correlation plot with strong correlation according to cell type origin. X indicates a non-significant correlation. Color intensity and the size of the dot are proportional to the Pearson correlation coefficient. Strong positive correlations are noted within cytokines linked with cytolytic T cell responses; macrophage responses; and T_H_2 responses. **(b)** Linkage clustering analysis of all samples demonstrates classes of samples based on the concentration of inflammatory cytokines. A minority of samples (grey class) had highest levels granzyme B, perforin, IL-6, IL-1α, MIP-1α and IFN*γ*. VL = viral load; DL = detection limit.

**Supplementary Figure 4:**
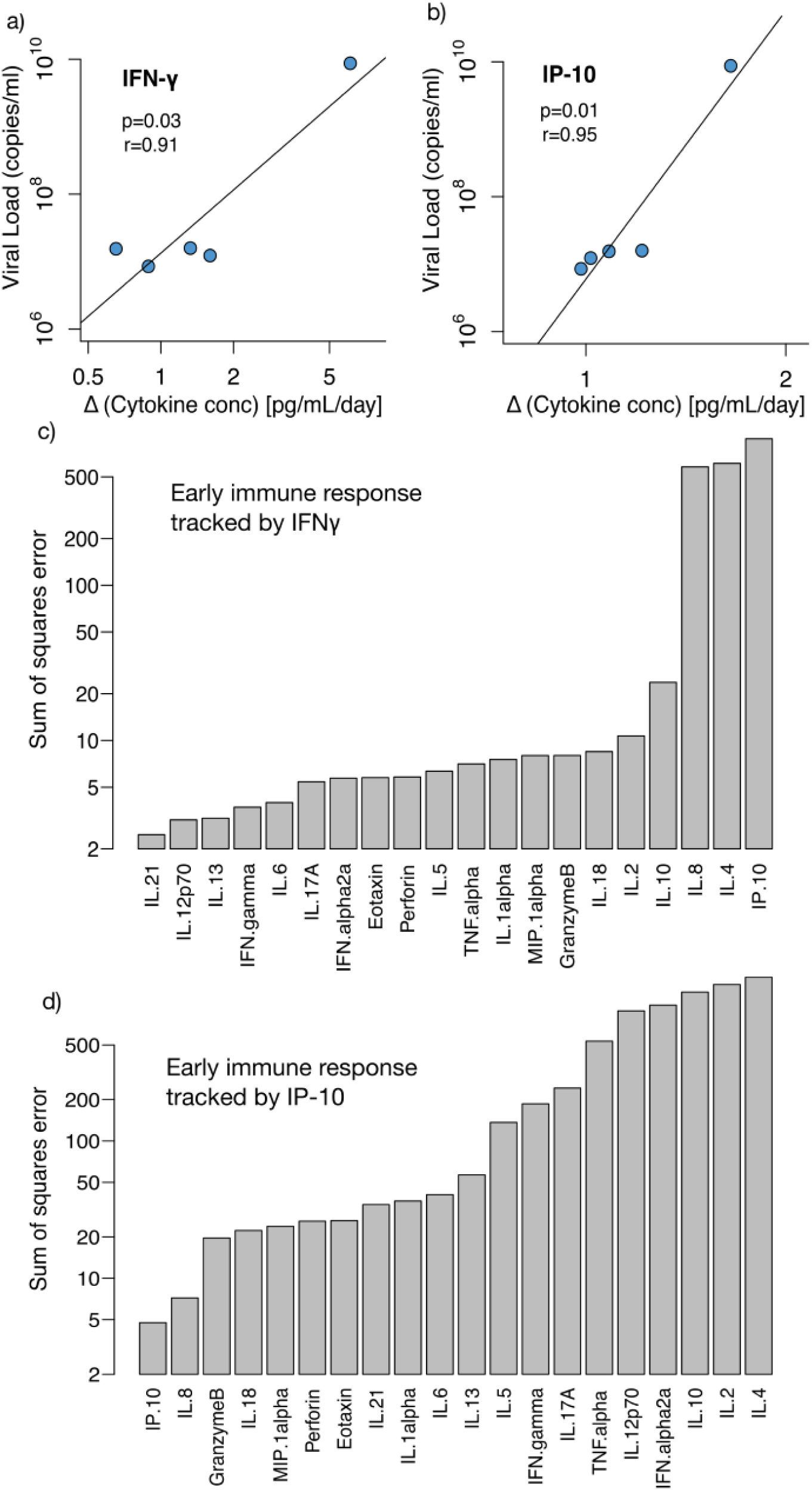
Selection of surrogate cytokines for modeling early and late immune responses against RSV. Scatterplot between RSV viral load from days 1 to 5 post enrollment and the log_10_ concentration of **(a)** IFNγ and **(b)** IP-10 until day 5 post enrollment. p-values and correlation coefficient obtained using Pearson’s test. Correlation test for all other cytokines was not statistically significant. **(c)** Sum of squared error (SSE) of the best model fits of equation (1) assuming early response (*G*) is tracked by IFNγ and late response (*C*) is tracked by each of the cytokines in x-axis. Lowest SSE (best fit) is obtained when late response is tracked by IL-21. **(d)** Sum of squared error (SSE) of the best model fits of equation (1) assuming early response (*G*) is tracked by IP10 and late response (*C*) is tracked by each of the cytokines in x-axis. Lowest SSE (best fit) is obtained when late response is tracked by IL-21.

**Supplementary Figure 5.**
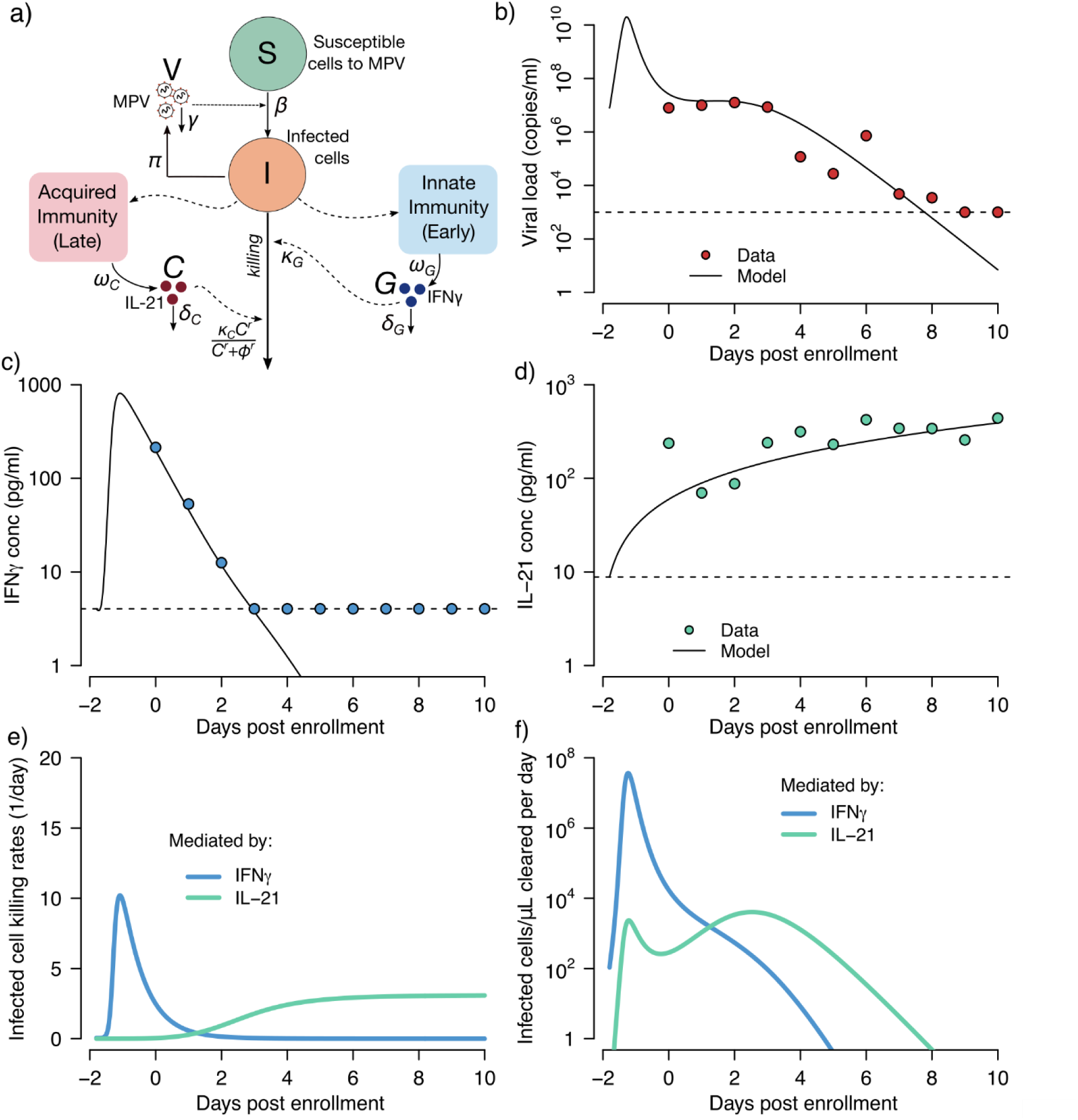
Mathematical modeling of a single participant’s MPV kinetics and the early and late immune responses tracked by IFN-γand IL-21. **(a)** Schematic representation of the model. *S* represents cells susceptible to MPV; *I*, MPV-infected cells; *V*, RSV virions; *G*, IFN-γconcentration and *C*, IL-21 concentration. Best fit models to **(b)** viral load, **(c)** IFN-γand **(d)** IL-21 measurements using a nonlinear least-squares approach. Circles represent the data, and black-solid lines the best model predictions. Models fit better to these cytokines than all others charted in **Fig 2** and **Sup fig 2. (e)** Model estimates of the killing rate per cell of infected cells mediated by IFN-γand IL-21, calculated as *κ* _*G*_ *G* and 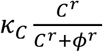,respectively. **(f)** Total number of infected cell deaths mediated by IFN-γand IL-21, computed as *κ* _*G*_ *GI* and 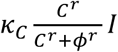, respectively. In **(e)** and **(f)** blue and green lines represent model predictions of the effects mediated by IFN-γand IL-21, respectively.

**Supplementary Table 1:** Symptom survey administered at enrollment (foam vs flocked swab comparison study) and daily (longitudinal sampling study).

| **Symptom Category** | **Specific symptom** |
| --- | --- |
| **Nose** | Runny nose |
|  | Congestion |
|  | Post-nasal drip |
|  | Sinus Pain |
|  | Sneezing |
| **Eyes** | Watery/burning eyes |
| **Ears** | Ear pain |
| **Throat** | Sore throat |
|  | Hoarseness |
| **Chest** | Cough |
|  | Phlegm production |
|  | Wheezing or chest tightness |
|  | Shortness of breath |
|  | Chest pain |
| **Gastrointestinal** | Diarrhea |
|  | Nausea |
|  | Stomach pain |
|  | Vomiting |
| **General** | Fatigue |
|  | Fever |
|  | Chills |
|  | Headache |
|  | Aching muscles |
| **Sleep Changes** | Sleep Disruption |
| **Sensory Changes** | Change in smell |
|  | Change in taste |

**Supplementary Table 2A:** Foam versus foam swab concordance in left nostril.

|  | **Flocked Swab** |  |  |  |
| --- | --- | --- | --- | --- |
| **Foam Swab** |  | **Positive** | **Negative** | **Total** |
|  | **Positive** | 4 | 2 | 6 |
|  | **Negative** | 2 | 7 | 9 |
|  | **Total** | 6 | 9 | 15 |

**Supplementary Table 2B:** Foam versus foam swab concordance in right nostril.

|  | **Flocked Swab** |  |  |  |
| --- | --- | --- | --- | --- |
| **Foam Swab** |  | **Positive** | **Negative** | **Total** |
|  | **Positive** | 5 | 3 | 8 |
|  | **Negative** | 1 | 6 | 7 |
|  | **Total** | 6 | 9 | 15 |

**Supplementary Table 2C:** Foam versus foam swab concordance with results from left and right nostril combined.

|  | **Flocked Swab** |  |  |  |
| --- | --- | --- | --- | --- |
| **Foam Swab** |  | **Positive** | **Negative** | **Total** |
|  | **Positive** | 9 | 5 | 14 |
|  | **Negative** | 3 | 13 | 16 |
|  | **Total** | 12 | 18 | 30 |

